# Predicting the impact of asymptomatic transmission, non-pharmaceutical intervention and testing on the spread of COVID19

**DOI:** 10.1101/2020.04.16.20068387

**Authors:** Ira B. Schwartz, James Kaufman, Kun Hu, Simone Bianco

## Abstract

We introduce a novel mathematical model to analyze the effect of removing non-pharmaceutical interventions on the spread of COVID19 as a function of disease testing rate. We find that relaxing interventions has a strong impact on the size of the epidemic peak as a function of intervention removal time. We show that it is essential for predictive models to explicitly capture transmission from asymptomatic carriers and important to obtain precise information on asymptomatic transmission by testing. The asymptomatic reservoir, reported to account for as much as 85% of transmission, will contribute to resurgence of the epidemic if public health interventions are removed too soon. Use of more basic models that fail to capture asymptomatic transmission can result in large errors in predicted clinical caseload or in fitted epidemiological parameters and, therefore, may be unreliable in estimating the risk of a second wave based on the timing of terminated interventions.

## I. INTRODUCTION

The ongoing Coronavirus Disease-19 (COVID19) pandemic, caused by the novel RNA virus SARS-CoV-2 [1], has demonstrated the need for swift and effective non-pharmaceutical intervention (NPI) strategies [2, 3]. In the absence of a cure or a vaccine and given the nature of the virus transmission, drastic measures need to be implemented to reduce contacts and “flatten” the epidemic curve. As quarantine and social distancing are enforced, it is important to understand the impact of early release or relaxation of such interventions on the affected population. We use a mathematical model which explicitly considers a NPI such as sheltering in place or social distancing, similar to what has been widely implemented in several countries around the world. We calculate the effect on the epidemic of removing intervention as a function of time. The use of drastic NPI strategies in China reportedly reduced the basic reproductive number, *R*_0_, to a value smaller than 1, strongly curbing the epidemic within a short period of time [2]. In other countries, the implementation of such policies has not been as strict [3], with an optimistic reduction in transmission by about half. To complicate the containment of the disease, early reports of pre-symptomatic and asymptomatic infections have emerged [4, 7], with estimates of asymptomatic transmission of as much as 85% of all cases, and 55% per person. These predictions have been supported by recent studies [18]. The US CDC estimates the population of asymptomatic infections to 25% of all cases [5].

The potential of COVID19 to spread unchecked puts a stronger pressure on the rapid design and deployment of accurate testing. Where rigorous and widespread testing protocols have been adopted, such as South Korea, the spread has been controlled [8]. An optimal control release strategy may include testing individuals before release. Feasibility of testing the whole population in a short time is under question. Therefore, it is important to understand how testing only a fraction of the asymptomatic population impacts the disease spread under different NPI scenarios.

Since public health interventions can affect symptomatic and asymptomatic individuals differently, we apply a model that explicitly captures both groups, as well as the case reporting rate which reflects the ratio of the symptomatic to asymptomatic infectious groups.

Transmission and recovery rates may differ between symptomatic and asymptomatic individuals. We compare the model and its predictions with the outcomes of a basic SEIR model (that fails to model asymptomatic individuals), which demonstrate that the basic model (SEIR) can not adequately predict the dynamics of Sars-CoV-2 (see Figure 16). For example, when the transmission from asymptomatic individuals differs from symptomatic ones, we show that applying a reproductive number that accounts for both groups leads to significant error in predicted clinical infectious cases by the simple SEIR model. Conversely, adjusting the SEIR model parameters to correctly account for the total reported cases forces significant error in *R*_*o*_ and incorrect timing for the epidemic wave. We report the relationship between the reproductive numbers in both models in the Supplementary section B.

As is common with emerging outbreaks of novel pathogens, the epidemiology of SARS-CoV-2 is not completely determined. Current estimations of the disease basic reproduction number, *R*_0_, put a realistic value between 2 and 3 [3, 6]. In our analysis, we use a baseline value of *R*_0_ = 2 (as a best case assumption). We also assume that implementations of control reduces transmission by 25% to 50%. The actual parameter values may differ and are still under investigations. However, the parameters we choose reflect a realistic scenario of transmission and are informative with regard to control strategies. Using these best case parameter, we then compute the additional cases that will occur as a function of the time when interventions are removed. The data suggests that decision to remove public health interventions can and should be informed by an analysis such as the one reported here. The fact that the epidemic curve is past its peak does not protect against a second wave so long as a large reservoir of unvaccinated susceptible population remains.

## II. MODEL DESCRIPTION AND RESULTS

To model the disease spread, we extend a classical Susceptible-Exposed-Infected-Recovered (SEIR) model to include an asymptomatic disease state, that is, a group of people capable of spreading the disease without showing any (or invisible) symptom which would induce testing. Asymptomatic transmission is thought to be mostly responsible for the world-wide distribution of the disease, as early travel bans imposed by China and other nations have focused on symptomatic cases (e.g., measuring body temperature at the airport) [6], thereby failing to control importation of the disease. Because of mild or no symptomatology, transmission from these asymptomatic carriers happens at a higher contact rate than symptomatic individuals. We parameterize these cases with the effective contact rate (transmission rate) *β*_2_, different from the transmission rate from symptomatic infectious, defined as *β*_1_. As such, susceptible individuals can become infected by coming in contact with both asymptomatic (*A*) and symptomatic infectious (*I*) individuals, which together compose the infected class. Individuals in the *I* compartment exhibit clinical symptoms. Individuals in the *A* compartment may not be included in case reports absent widespread testing. We assume a proportionality constant (related to the case reporting rate) regulating the relative rate at which exposed individuals (who have come in contact with the disease but are not yet infectious), contribute to the *A* and *I* compartments respectively.

With the hypothesis that social distancing reduces transmission by 25% and considering a 25% fraction of asymptomatic infectious, it is possible to see a two-fold reduction in the peak proportion of infected (Figure 1). If control is released before the epidemic peak, at *t* = 0.28 years, the model predicts a strong resurgence of the disease, with no flattening and an infectious fraction almost reaching the uncontrolled epidemic. Of course, while a delay in the peak is observed, which may buy the health care system time to respond, the reduction in the number of total infectious is marginal. When NPI is removed at the epidemic peak (figure 1(b)), a secondary peak is observed, showing an increase in total number of infected. Lastly, when control is released after the peak (at a late stage of epidemic shown in figure 1(c)), for *t* ≈ 0.5 years, we observe the emergence of a kink in the descending part of the curve, but not a full fledged resurgence of the disease.

**Figure 1:**
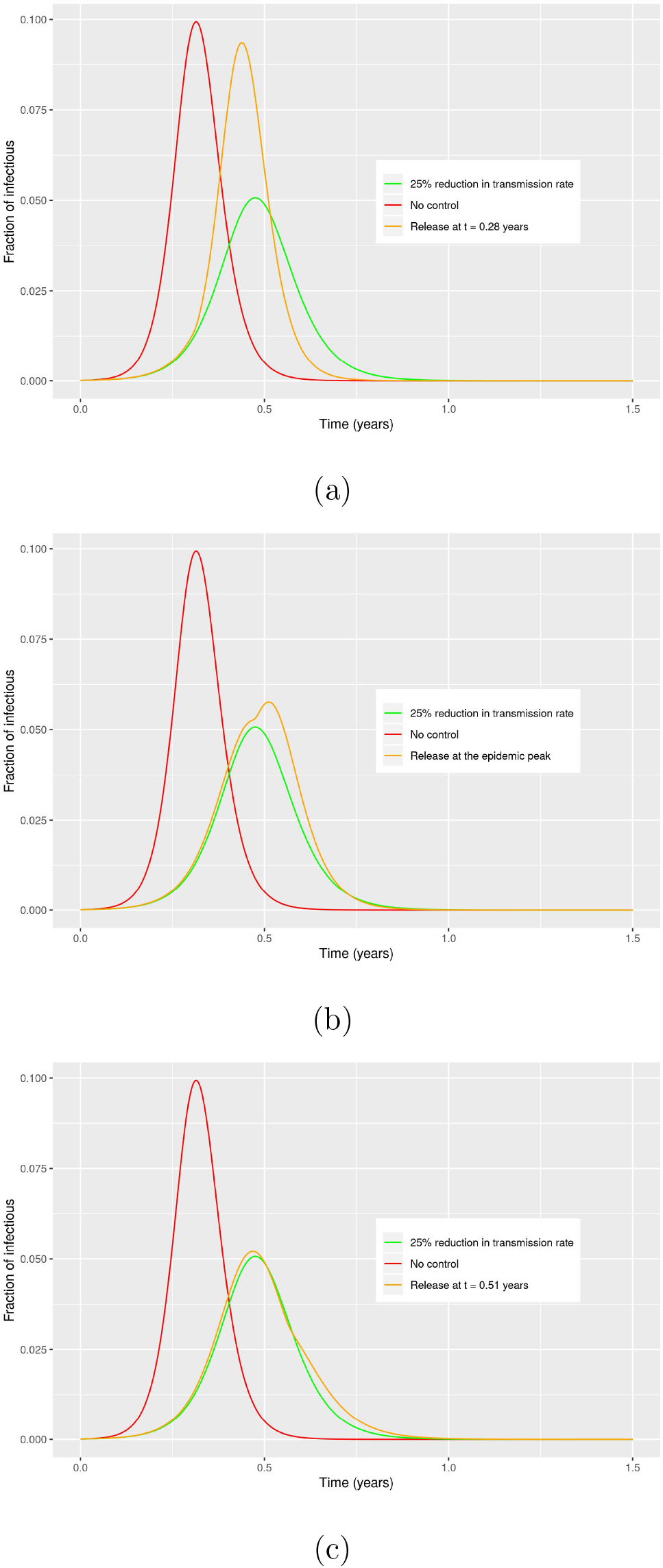
Effect of early removal of non medical intervention on the dynamic of the disease. The red curve shows the unmitigated epidemic, i.e. no intervention. The green curve is the effect of reducing transmission rate by 25% as consequence of reducing social interactions. The orange curves show release of control at various stages of the epidemic curve: (a) at *t* = 0.28 years, (b) at the epidemic peak, and (c) at *t* = 0.51 years. Asymptomatic infectious are 25% of all infectious cases.

The outbreak has a very different shape when infectious include a large proportion of asymptomatic individuals (Figure 2). In this case, early release of intervention causes a dramatic resurgence of the total infections, even for stronger control and at a late stage of the epidemic.

**Figure 2:**
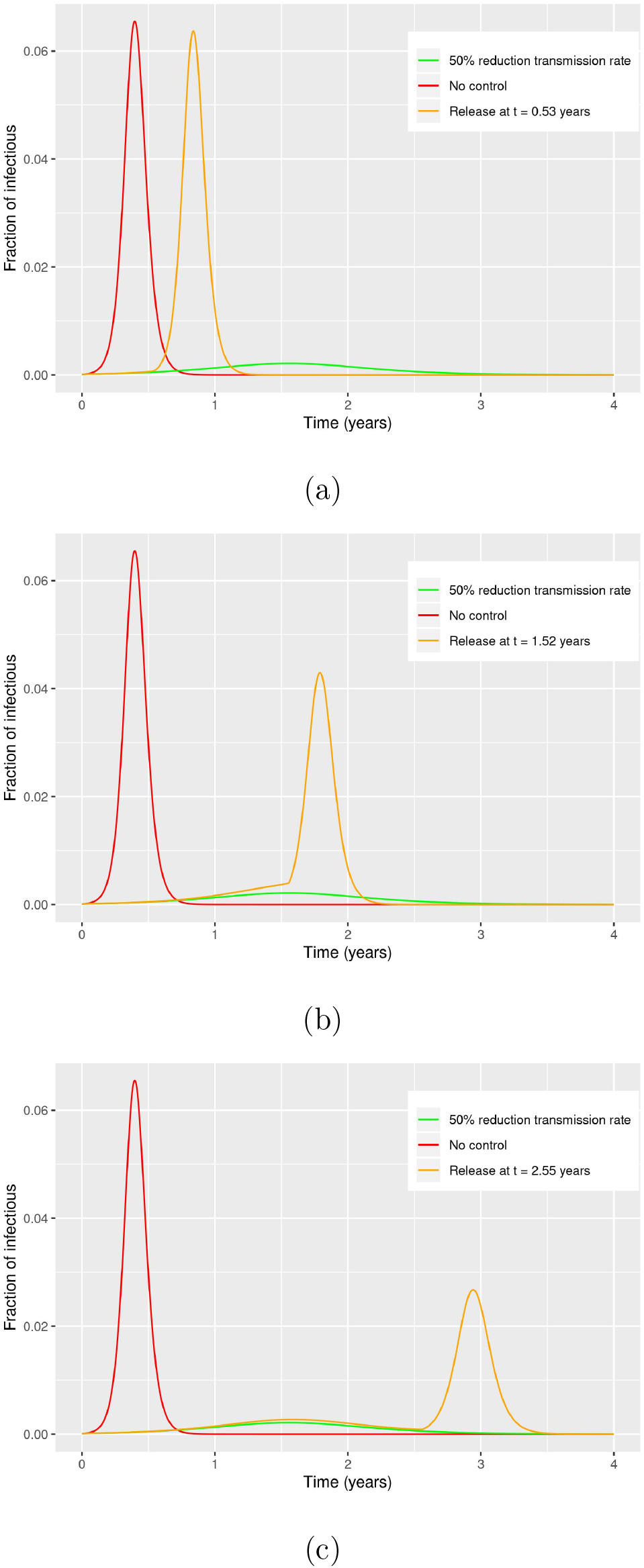
Effect of early removal of non medical intervention on the dynamic of the disease. The red curve shows the unmitigated epidemic, i.e. no intervention. The green curve is the effect of reducing transmission rate by 50% as consequence of reducing social interactions. The orange curves show release of control at various stages of the epidemic curve: (a) at *t* ≈ 0.5 years, (b) at the epidemic peak, and (c) at *t* = 2.5 years. Asymptomatic infectious are now 85% of all infectious cases.

When evaluating the effect of NPIs, we calculate the total cumulative incidence (including asymptomatics) caused by releasing intervention for a sample population of 100000 people (Figures 3 and 4). Simulations show that early release of intervention causes an expected total number of cases similar to the case of no intervention. If one considers asymptomatics to make for 1 in 4 people as suggested by the CDC, release at later time show a delay in the peak and a reduction in cumulative incidence between ≈ 10% and 20%, which translates into ≈ 5000 to 10000 less infected people. Even a conservative estimation of the disease death rate results in a large impact of different policies on the population. On the other hand, when asymptomatics make for the majority of the infections, as recent data seems to suggest, the release of NPI even after a long time is predicted to cause a strong resurgence and a large impact, as measured by the cumulative incidence, and a large second wave of infection is observed. When taken together, the results in Figures 1 through 4 show the need to obtain precise information about the fraction of the population which is actually currently infected with the disease before any release of control can be decided.

**Figure 3:**
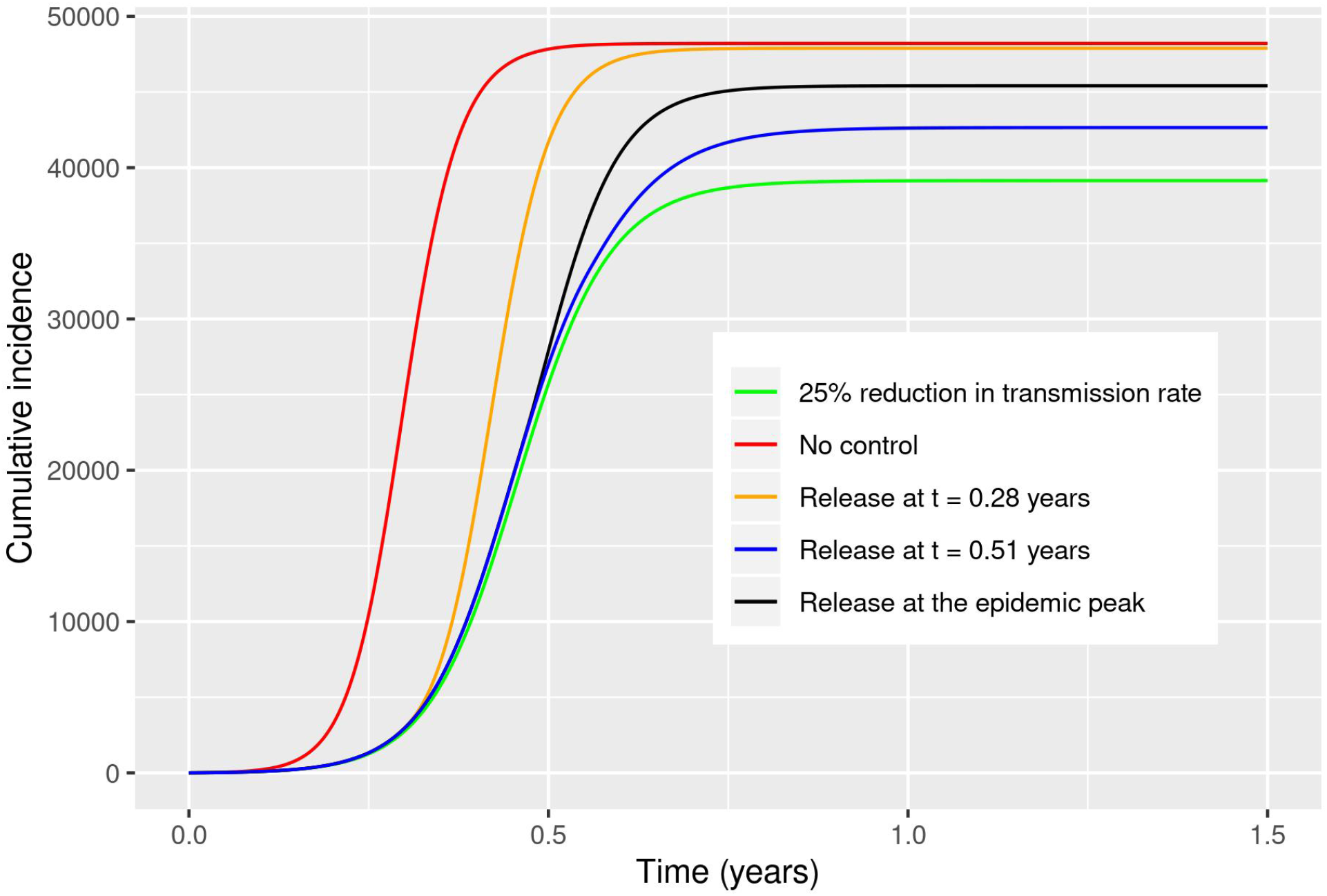
Cumulative incidence for an example population of 100*k*, with release of control at different times. Asymptomatics make for 85% of the infectious population. 25% reduction of transmission is achieved by reducing the asymptomatic transmission rate.

**Figure 4:**
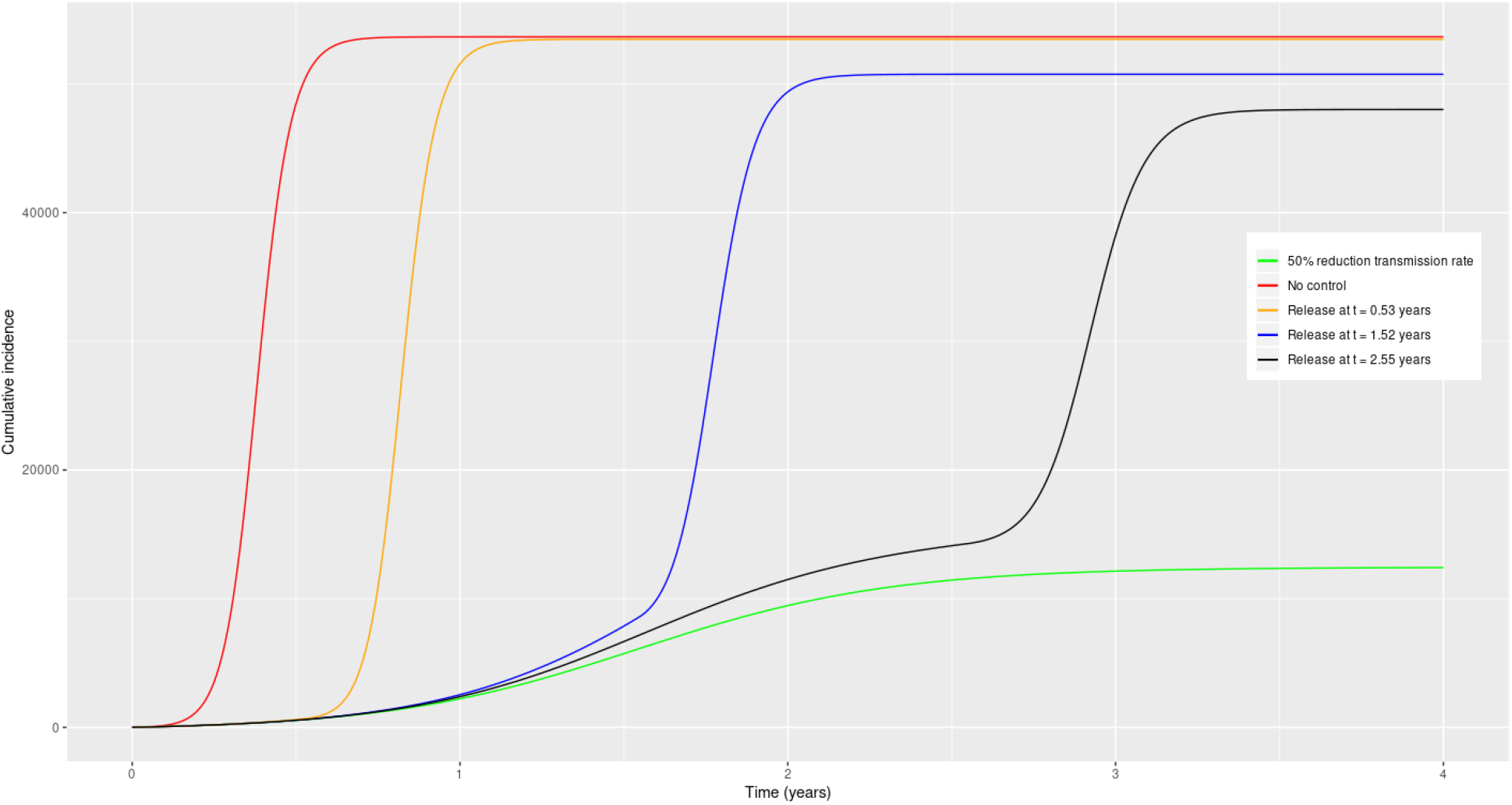
Cumulative incidence for an example population of 100*k*, with release of control at different times. Asymptomatics make for 85% of the infectious population. 50% reduction of transmission is achieved by reducing the asymptomatic transmission rate.

## III TESTING AND SOCIAL DISTANCING

We next extend of our model to describe the impact that testing a fraction of the population would have on the disease dynamic. We assume that testing captures only a part of the asymptomatic population, indicated as *A*_1_. Testing susceptible and recovered has no effect on the epidemic curve, but detecting asymptomatic cases requires significant random sampling. We further assume that widespread testing effectively removes *A*_1_ asymptomatic individuals from the population transmitting the disease through self isolation or quarantine. We model this removed *A*_1_ population fraction by assuming an increased recovery rate for this class (since isolation is akin to removal). A fraction *A*_2_ is not tested and will be able to spread the virus upon release of NPI as before. It is informative to consider the case of testing both with and without social distancing. We imagine two scenarios: In the first scenario, as soon as the first confirmed case is reported, a fraction of the population is tested but no social distancing is implemented. This is reported in Figures 5 and 7. The cumulative incidence for this scenario is shown in Figures 6 and 8 for 25% and 85% fraction of asymptomatics, respectively. Our simulations show that, regardless of the composition of the infected class, a secondary peak of infection is not observed when testing is implemented on a fraction of the population. However, an early testing policy that catches even 25% of all asymptomatic cases results in a significant reduction of the number of new cases per 100*k* (≈ 1000 less cases predicted per 25% of the population tested).

**Figure 5:**
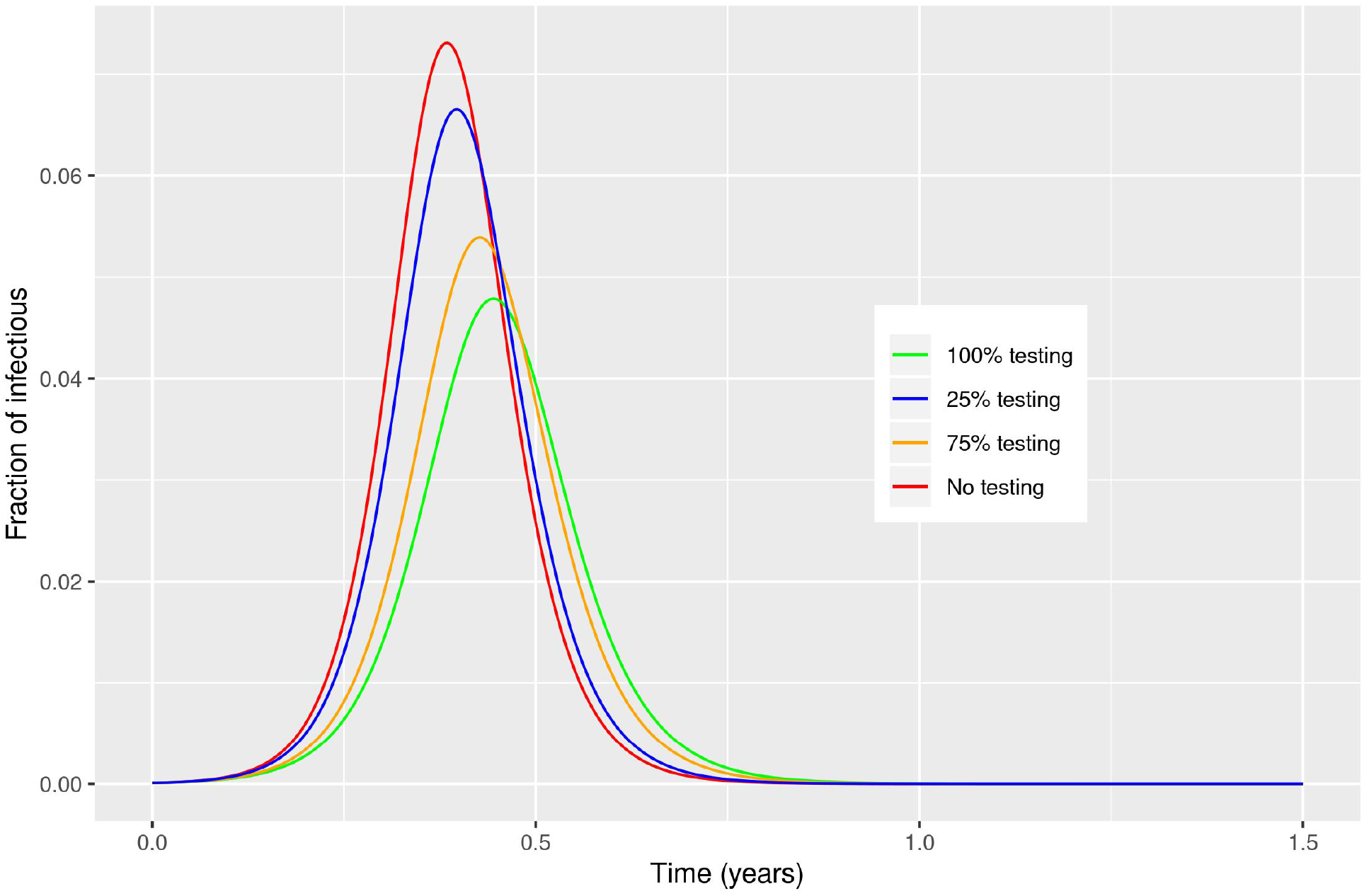
Effect of testing a fraction of the asymptomatic population as soon as the first case of infection is reported, in the absence of additional NPI. The asymptomatic make for 25% of all infectious cases. While a secondary peak is not observed, testing asymptomatics is predicted to reduce the intensity of the epidemic peak.

**Figure 6:**
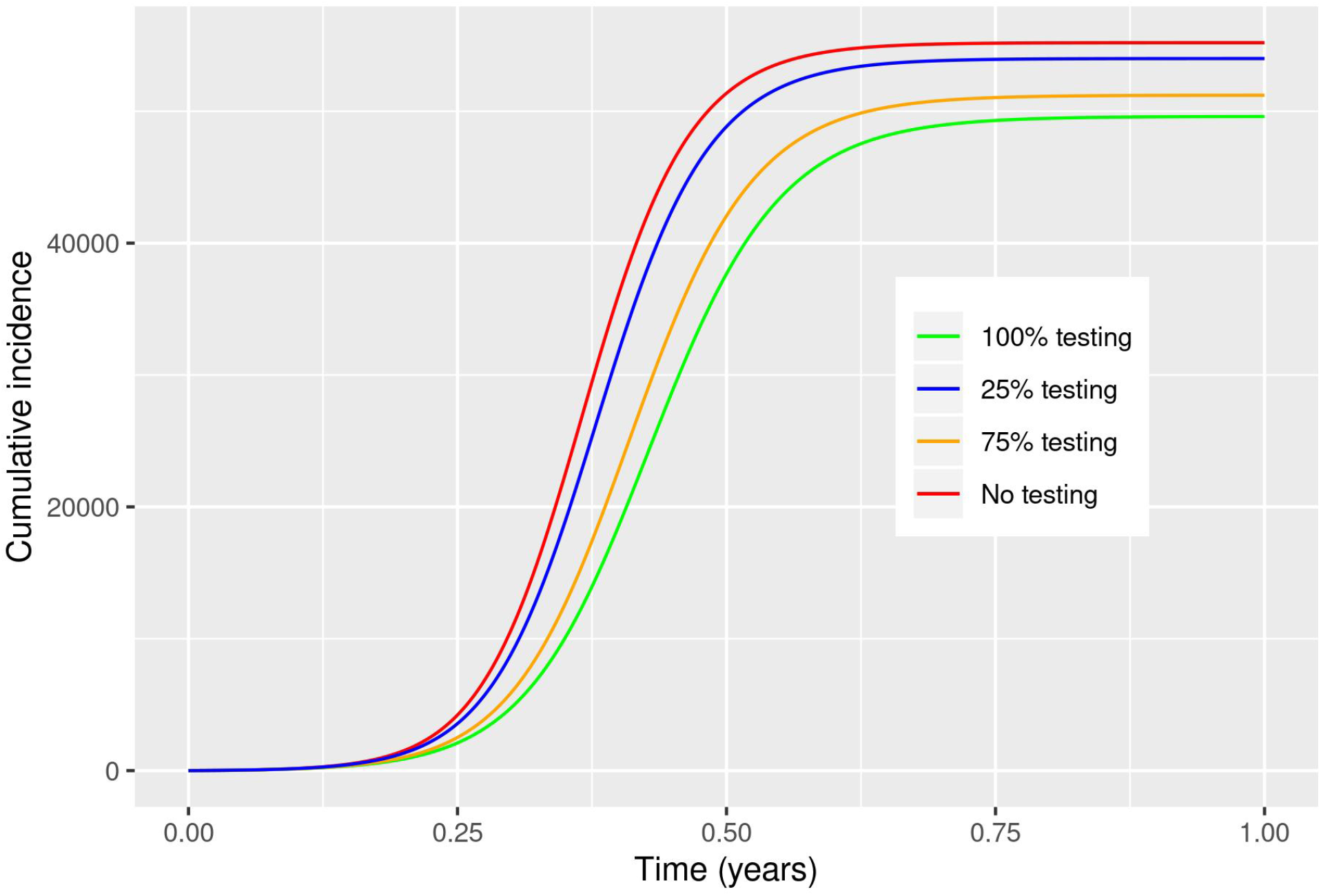
Cumulative incidence for a example population of 100*k* people, for various fractions of the asymptomatic population tested. Asymptomatics are 25% of the infectious population.

**Figure 7:**
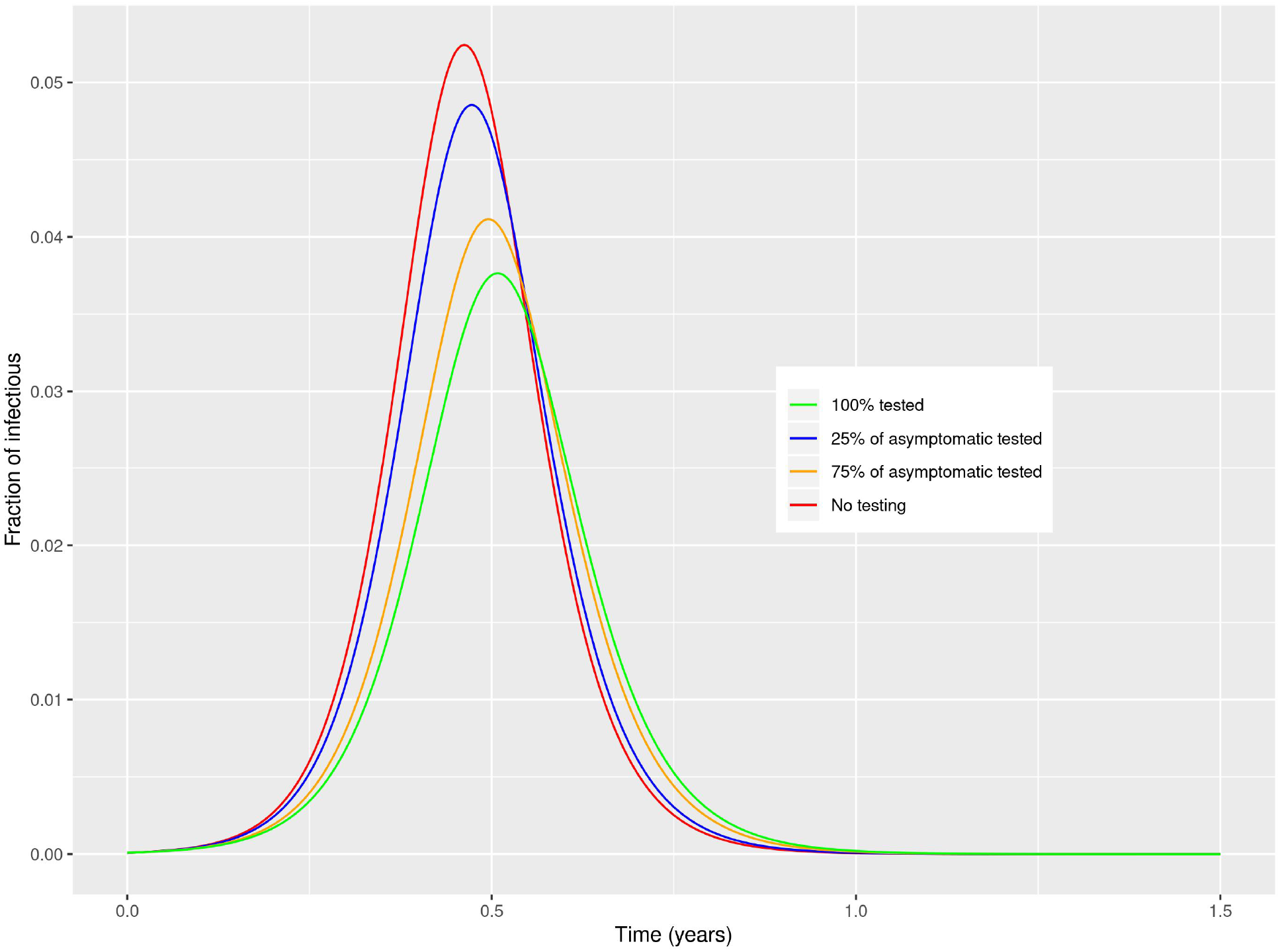
Effect of testing a fraction of the asymptomatic population as soon as the first case of infection is reported, in the absence of additional NPI. The asymptomatics make for 85% of all infectious cases. While a secondary peak is not observed, testing asymptomatics is predicted to reduce the intensity of the epidemic peak.

**Figure 8:**
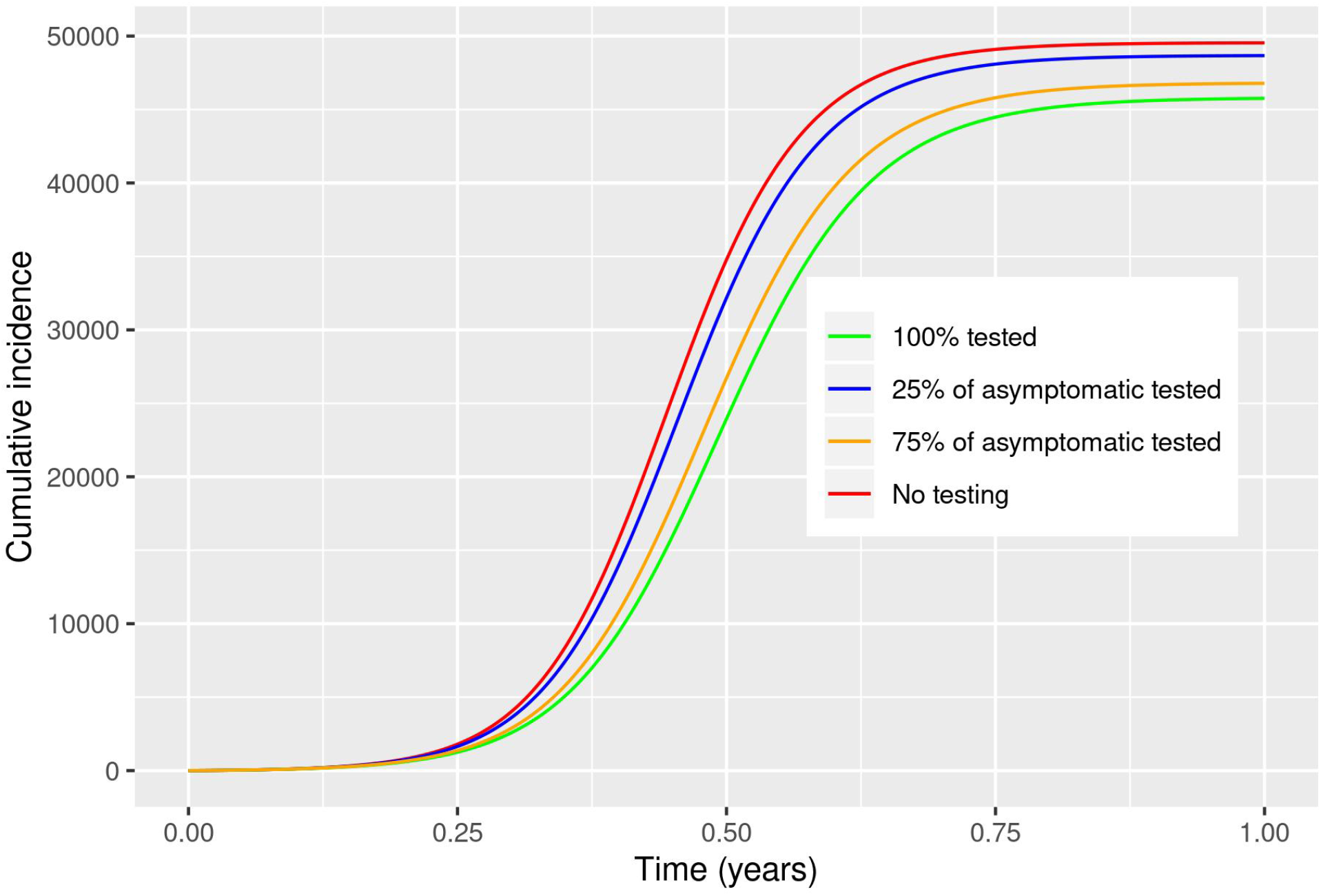
Cumulative incidence for a example population of 100*k* people, for various fractions of the asymptomatic population tested. Asymptomatics are 85% of the infectious population

In the second scenario, we consider relaxing NPI only after testing a fraction of the population. This is reported in Figures 9 through 12. In this scenario, we implement NPI when the first case is reported. Testing serves as a mean to relax NPI by releasing the untested fraction of the population to their dynamic in the absence of social distancing. Again, asymptomatic individuals that are detected through testing will be removed from the population at a higher rate, while asymptomatic untested are allowed to transmit the disease without restrictions. If the asymptomatic carriers are 25% of the total infected population, removing NPI before the epidemic peak when testing a fraction of the population (e.g. 25%) results in a 5 to 10% increase in the disease burden. Testing at or after the peak results in an epidemic curve which closely follows the full control. However, when the fraction of asymptomatics makes for the majority of infected cases, the results are markedly different. Even if NPIs are relaxed at a later time, a strong resurgence of the epidemic is observed. The presence of a large fraction of asymptomatics that resume social activities untested is predicted to cause ≈ 30000 (30%) more cases than the full control, even if 25% of all asymptomatic are found through testing.

**Figure 9:**
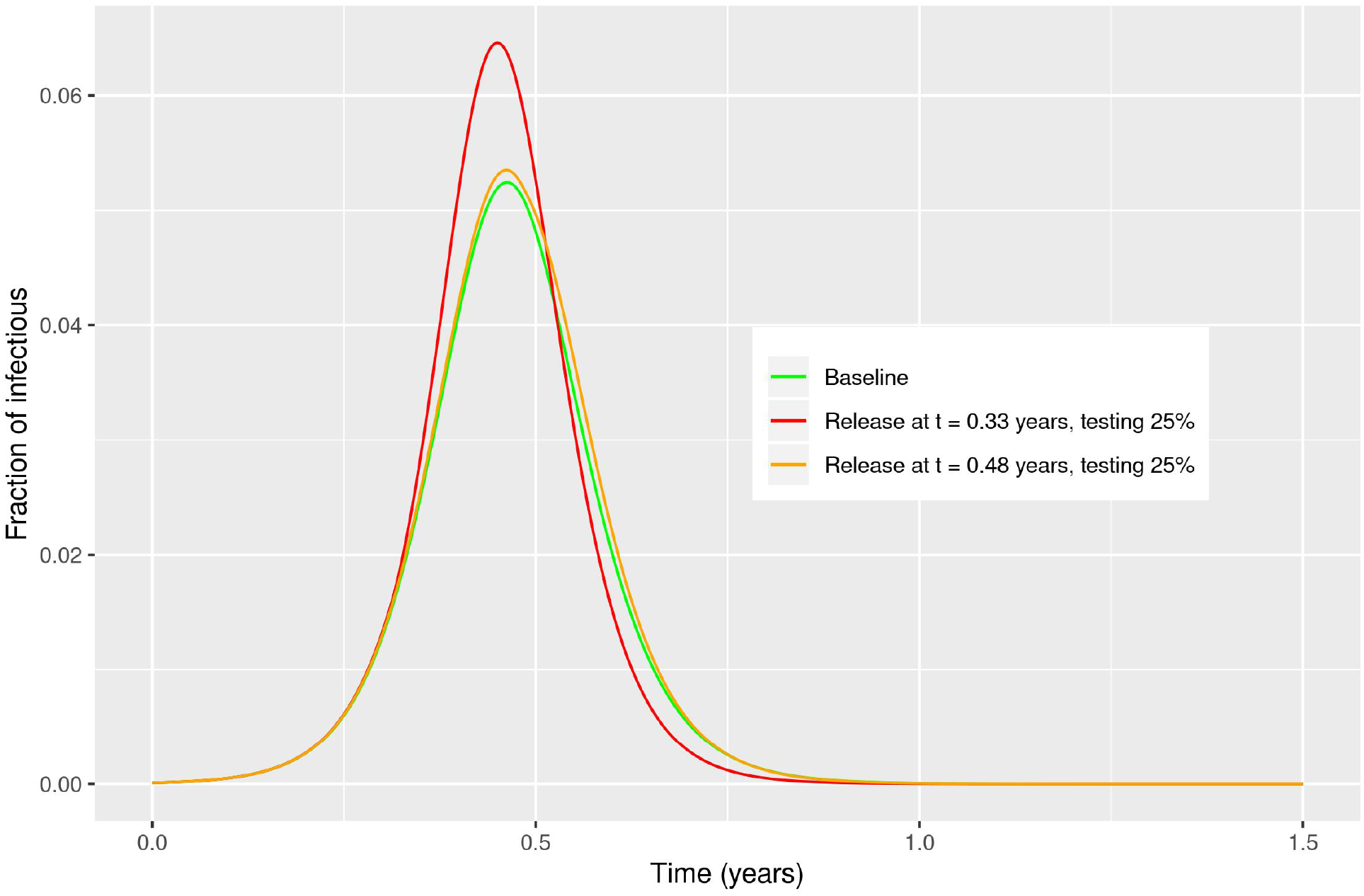
Effect of testing and social distancing, at different times, when asymptomatics are 25% of the infectious population. If 25% of the population is tested, a strong resurgence of the epidemic is observed if release of NPI occurs before the epidemic peak. On the other hand, testing at or after the epidemic peak results in a negligible difference between full control and release of control.

In conclusion, we have described the combined effect of release NPI and testing strategies as applied to the current COVID19 epidemic. Our simulations clearly show that, absent testing of a significant fraction of the population (both symptomatic and asymptomatic), early release of NPI will cause a resurgence of the epidemic. This suggests the fundamental role of widespread testing in any policy of NPI release. As suggested by recent publications [18], when universal testing is administered, asymptomatic individuals are found to make up for a large majority of all identified cases. As such, widespread testing policies must target this population rather than only individuals with evidence of influenza-like or SARS-like symptoms.

As a final remark pointing to further study, quarantines such as those who are performing self-imposed isolation, are similar to the case where people change their behavior in the presence of a fatal disease. Adjusting their behavior to avoid those who are contagious leads to a dynamic network where the connections depend on the state of the population of individuals. That is, the network of contacts is one that is adaptive [11]. Although there does not exist a vaccine currently, when one will be introduced, it will initially be a limited resource. Continuing controls, such as those adaptive contacts in which people can avoid contagious individuals, can greatly help with the extinction of disease [12], especially in the presence of limited vaccines. Combining large scale testing, continued social distancing, along with vaccine control increases the probability of being able to bring the COVID19 disease to extinction in the presence of limited resources.

## Data Availability

No relevant data is present in the article

## IV. ACKNOWLEDGEMENTS

IBS acknowledges useful discussions of the modeling with Janine Tucker of McKean Defense, as well as support from the NRL Base funding no. N0001420WX00410, as well as the Office of Naval Research no. N00001419WX01322. KH, JHK and SB acknowledge discussions with the IBM COVID19 modeling taskforce. SB also acknowledges discussions with the Cellular Engineering team at IBM Research Almaden.

## Appendix A: Equations for the SEAIR model

The equations for the SEAIR model are:

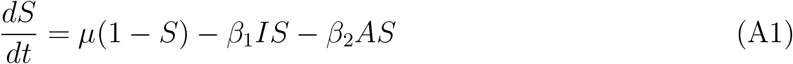

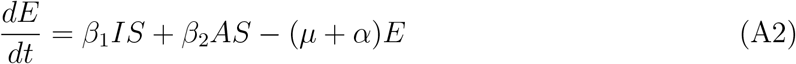

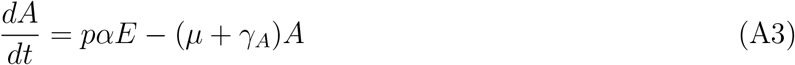

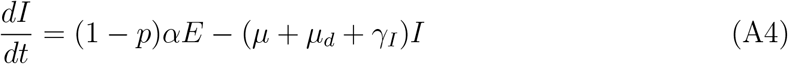

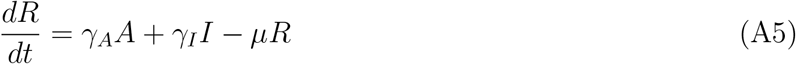

We indicate with (*β*_1_, *γ*_*I*_), (*β*_2_, *γ*_*A*_), respectively, the symptomatic and asymptomatic transmission and recovery rates, *α* is the incubation rate, *p* is the fraction of asymptomatic infectious. We assume also that *µ*_*d*_ is the case mortality rate due to the virus for symptomatic individuals, but not asymptomatics. It is different from the background birth and death rate, *µ*.

To find the fraction of susceptibles in the population and thus *R*_0_, we let *κ* be function of *p*:

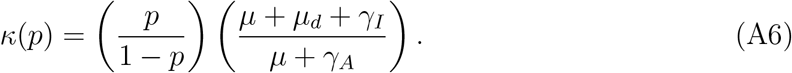

From the equilibrium state, we find the following reproductive numbers in terms of the rate parameters:

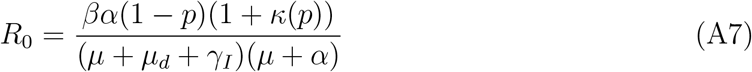

Notice that in the limit of 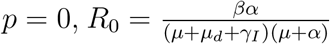, which is the reproductive rate for the SEIR model without asymptomatic individuals. NPIs and release thereof are implemented as difference in the contact rates, *β*_1_ and *β*_2_. When control is implemented, then *β*_1_ = *β*_2_ and lowered to reflect social distancing. We report the overall *R*_0_ values for the different contact rates under control and release.

## Appendix B: SEAIR with testing and social distancing

We assume that testing asymptomatic individuals has the effect of increasing the recovery rate so that tested asymptomatic individuals are removed from the population at a faster rate. The equations read:

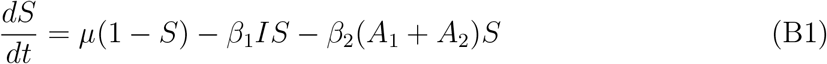

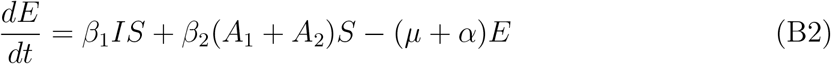

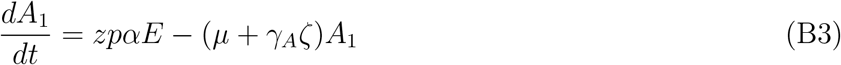

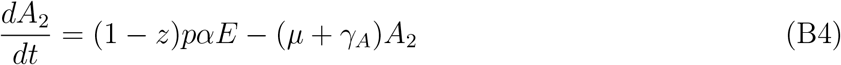

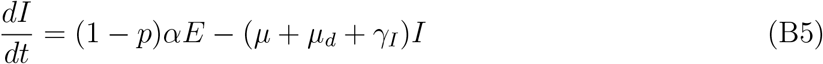

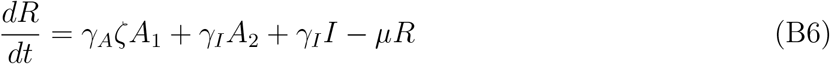

where *z* ∈ [0, 1] represents the fraction of A that are tested, and *ζ* ≥ 1 is the increase in the rate of recovery for those tested, since they are removed from the population at a faster rate. Note that since the the recovery rate of tested asymptomatics decreases the fraction of total *A* = *A*_1_ + *A*_2_, the effective contact rate between *S* and *I* is already reduced.

We can rederive the reproductive rate of infection as before, by expressing the steady state epidemic solution all in terms of *I*. The result is then given by

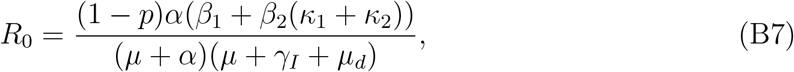

where

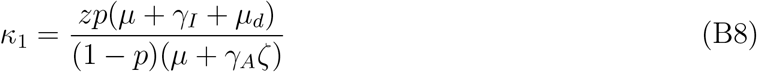

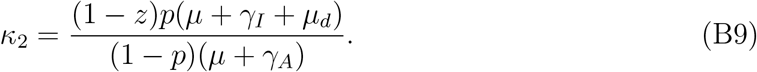

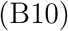

Assuming *z* = 0, *ζ* = 1; i.e., no testing, we have

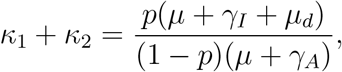

and

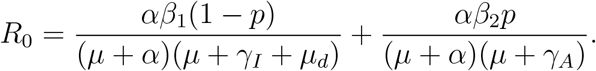

Limiting cases:

1. *P =* 0 implies 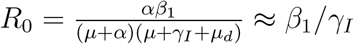
2. *P =* 1 implies 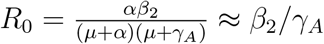

We remark that for Figs. 9 and 11, we chose the contact rates so that *R*_0_ = 1.77, 1.11 respectively, and assumed that the contact rates were chosen so *β*_2_ = *β*_1_/2.

**Figure 10:**
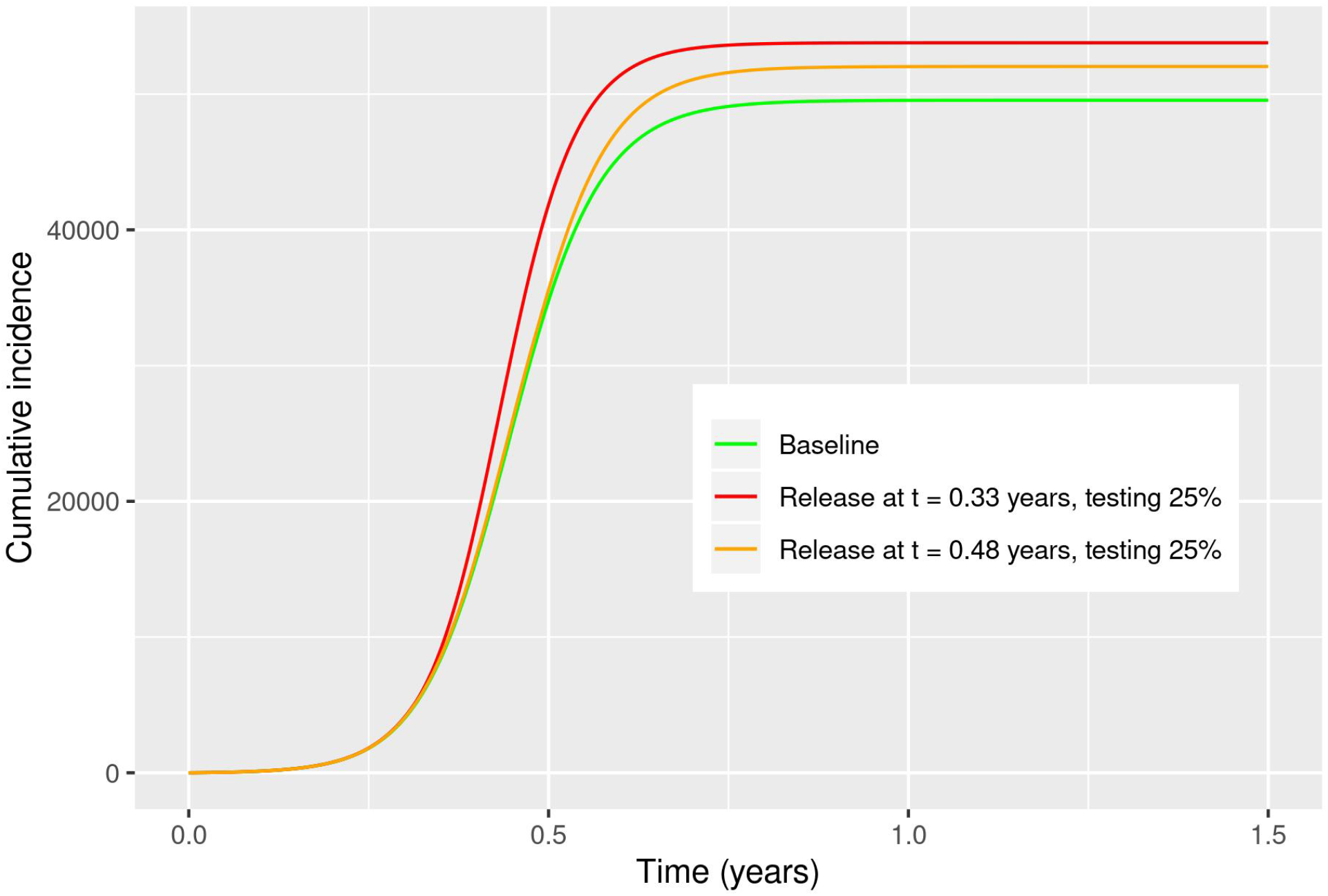
Cumulative incidence for a example population of 100*k* people. NPI are assumed in place, and release is assumed at time of testing. Asymptomatics are 25% of the infectious population.

**Figure 11:**
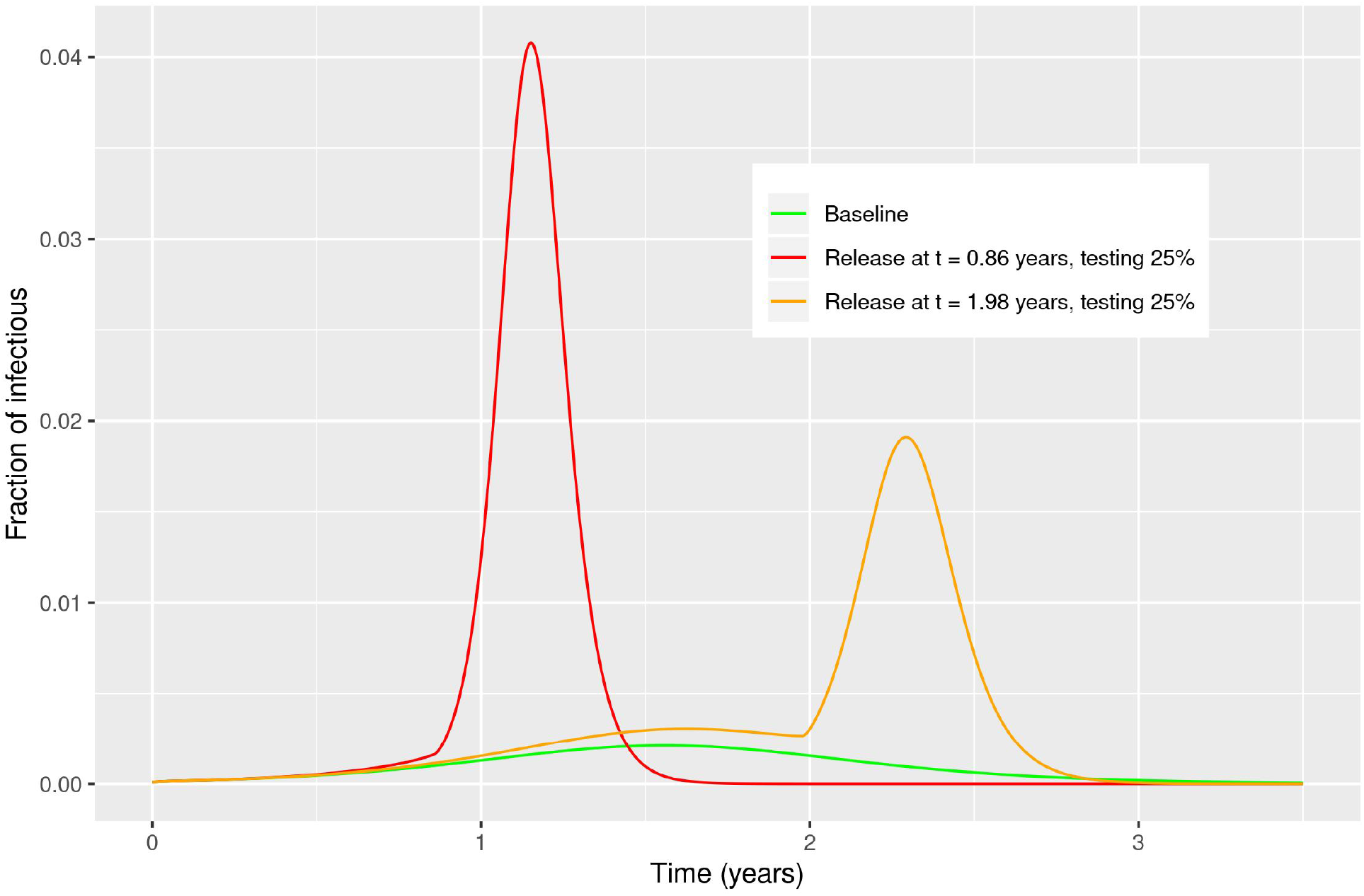
Effect of testing and social distancing, at different times, when asymptomatics are 85% of the infectious population. If 25% of the population is tested, a strong resurgence of the epidemic is observed if release of NPI occurs before the epidemic peak. Testing at or after the epidemic peak results in a strong secondary peak of infection.

## Appendix C: SEIR model structure

For the SEIR model with infectious deaths, we assume the following:

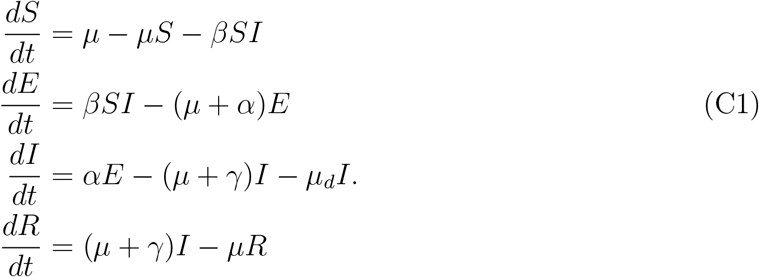

Notice that in the infected equation, there is an extra death term, *µ*_*d*_. The idea is to incorporate the deaths due to infection independent to regular birth and death rates.

Using this model, the reproductive rate of infection becomes

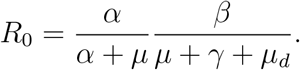

With the extra removal of infected by death, the contact rate, *β*, must be increased to sustain a given value of *R*_0._ For example, with *µ*_*d*_ = 0.035/year, and the parameters given by *µ* = 0.02, *α* = 36.5, *β* = 36.5, we find that to achieve an *R*_0_ = 2 requires *β* ≈ 73.1/year.

In Figure 13, we consider the case where the population has effective contact rates on average that yield a reproductive rate of infection, *R*_0_ = 2. We plot the predicted infected and exposed fractions of the population as a function of time in years. Starting from a very initial infected, we that the peak is reach in approximately 0.5 years, or 26 weeks. In addition, for such a low reproductive rate, the exposed class precedes the infected with a slightly higher peak.

**Figure 12:**
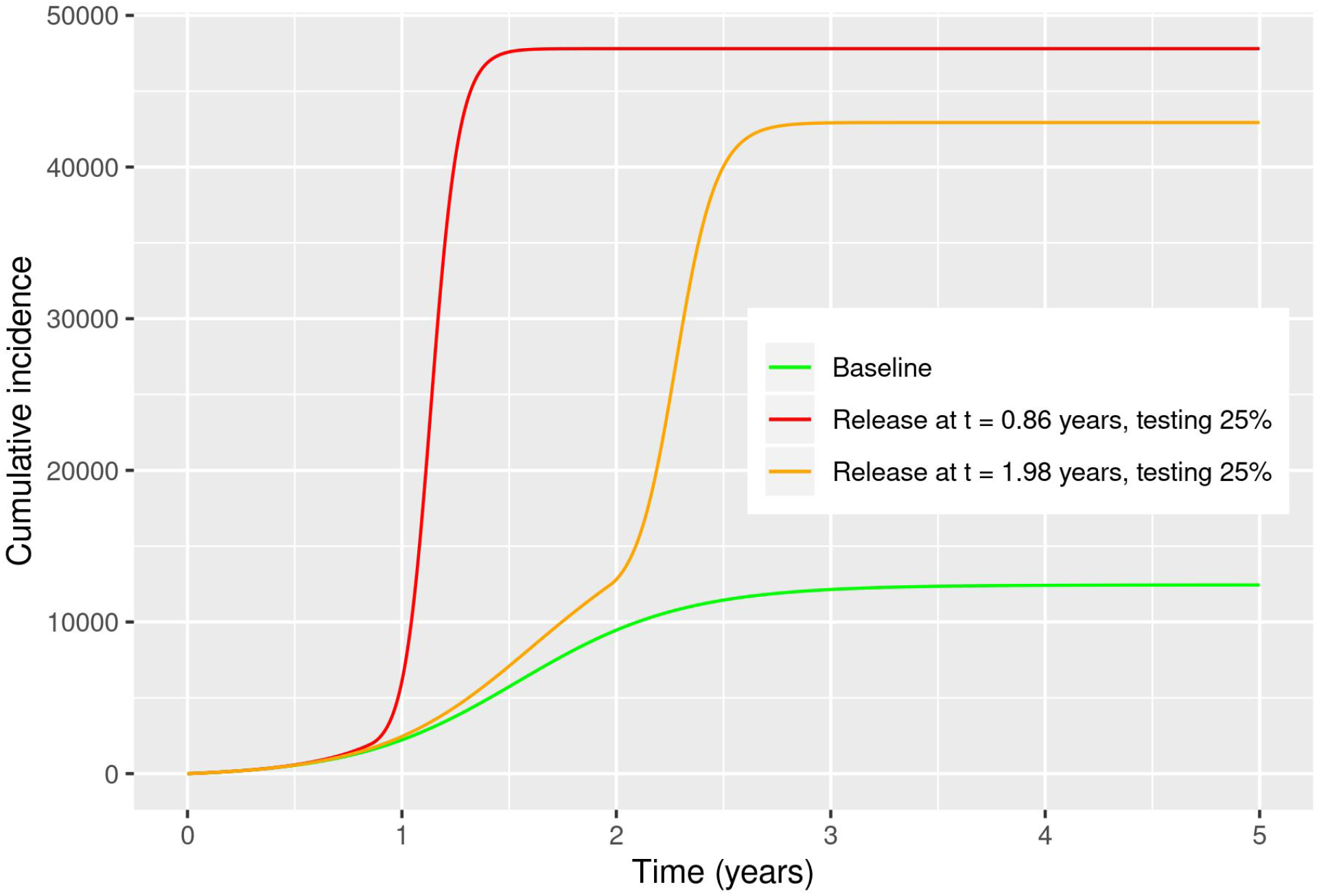
Cumulative incidence for a example population of 100*k* people. NPI are assumed in place, and release is assumed at time of testing. Asymptomatics are 85% of the infectious population.

**Figure 13:**
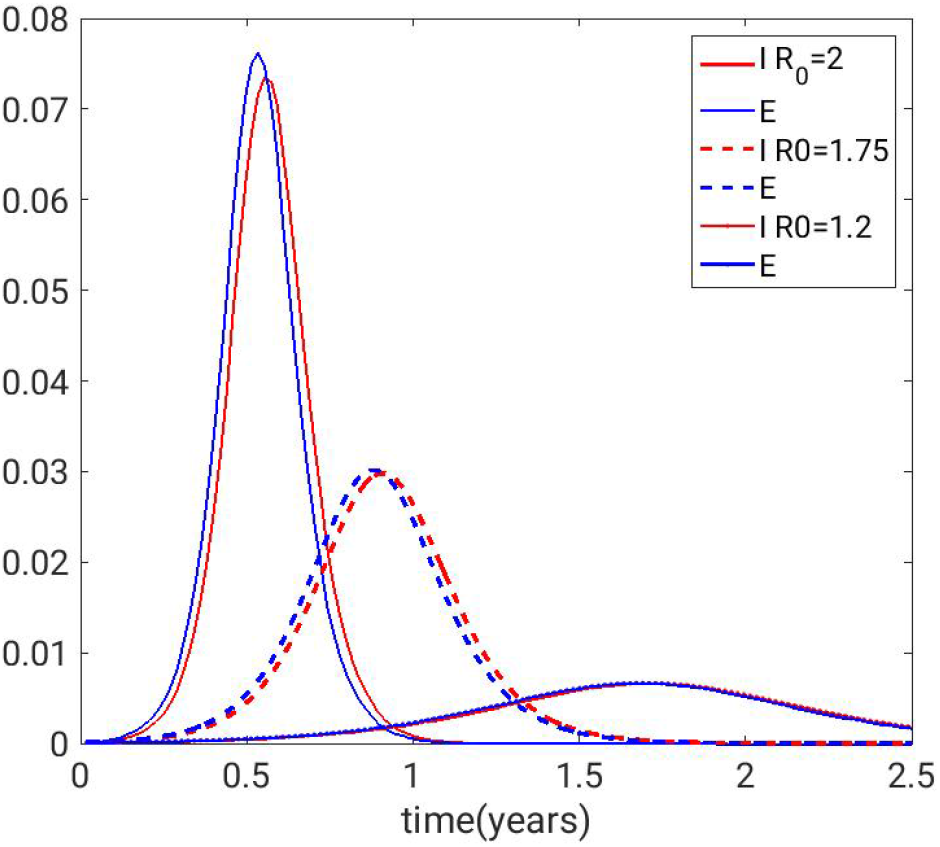
Exposed (blue) and infected (red) for *R*0 = 2, 1.75, and 1.2. *R*0 = 1.75 corresponds to quarantining 1 in 4 people from the population. That is reflected in a 25 percent reduction in the contact rates. Other parameters where rates are in years: *µ* = 0.02, *µ*_*d*_ = 0.035, *α* = *γ* = 36.5. *β* is adjusted to get the appropriate value of *R*_0_.

We then assume that controls are induced by requiring the population to remove 1 in 4 people, or reducing the effective contact rate by 25 percent. The results in Figure 13 are shown for both *R*_0_ = 1.75, 1.2. In both cases, as expected the peak is reduced greatly, and shifted in time. The implication is that even with constant control of quarantine, the virus persists. In order to achieve an *R*_0_ = 1, which is needed drive the disease to extinction, *β <* 36.55/year for the chosen demographic and epidemic parameters.

We now consider interventions which are phased depending on the population size, and whether the incidence is increasing or decreasing. The results are plotted in Figure 15. The relative peaks on the upward slope when controls are released clearly explode. Even if controls are released at the peak of the steady control curve, there is an increase in the fraction of infectious. A similar behavior is observed if control is released at later times.

**Figure 15:**
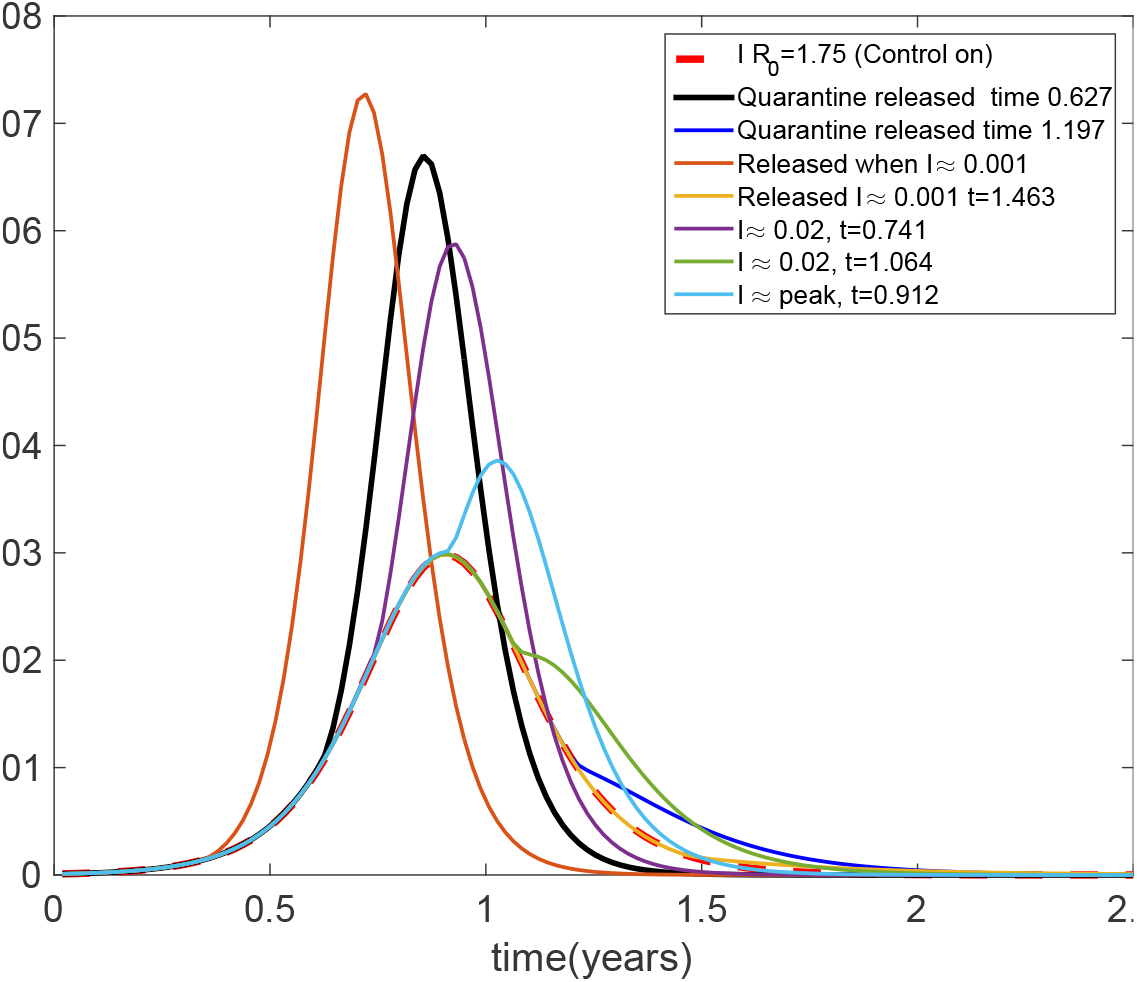
NPIs are released during control measurements. The red dashed line depicts the results for continuous control for all time. The other curves release control at various times on the upward and downward slopes of the control curve (red dashed line).

## Appendix D: Effect of Non-pharmaceutical Intervention relaxation on the SEIR model

Since we see a definitive effect of NPI on the population for various fractions of the population, we consider here how the effects of turning off controls by allowing people to remix with the population; i.e., what happens when control is turned off. To quantify the effects of controls release, we first plot the case where there is no control, so we can compare re-ignition peaks, and also the case where 1 in 4 (25 per cent) are quarantined (i.e., NPI intervention is implemented). In Fig. 14, they are plotted as solid and dashed red lines, respectively. Notice that with controls always on, the peak of the epidemic is reduced by about 2.5 times.

**Figure 14:**
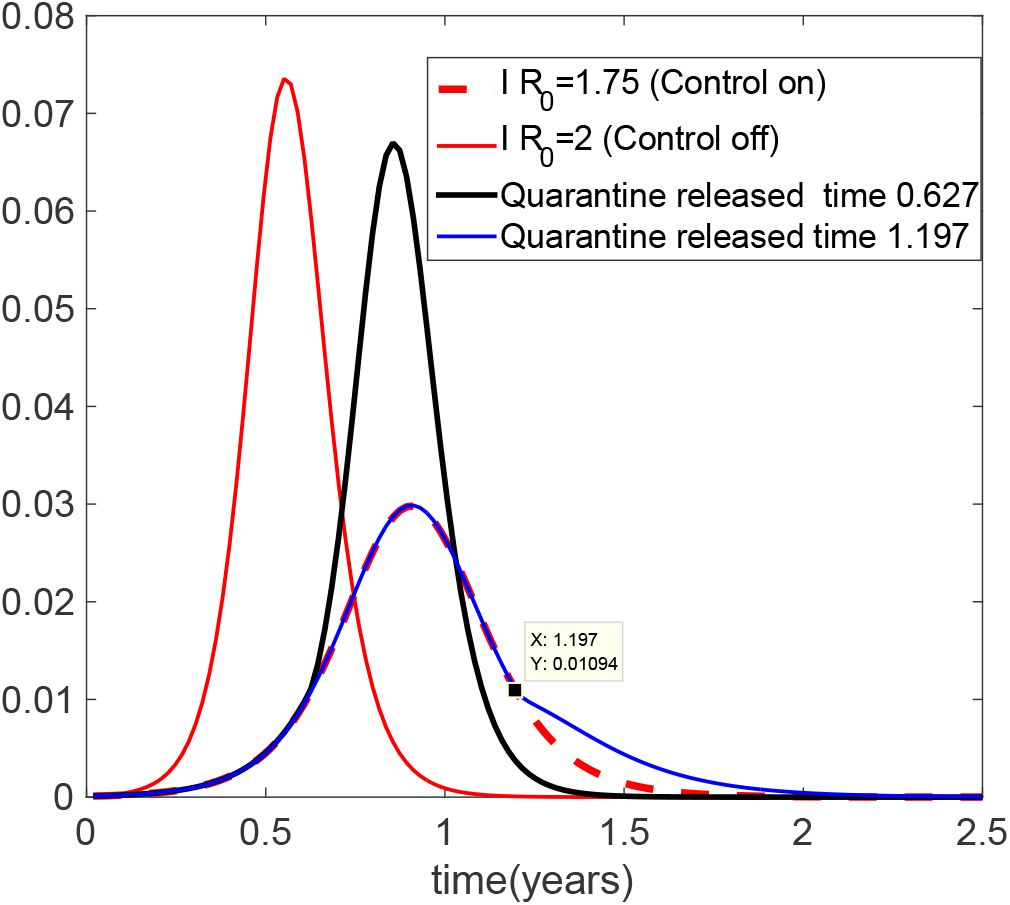
Depicted shows the uncontrolled infected (Solid red line), the controlled infected (dashed red) when *R*_0_ = 1.75. Two different controls are implemented when the infected fraction hits approximately 1 per cent. The black line releases the controls on the upward slope, while the blue line turns off control on the downward slope.

We now consider the same controls, but we turn off quarantine when the fraction of infected population is approximately 0.1, or 10 per cent. In the first case, we release quarantine, on the upward slope of the infected population. The results are shown as a solid black line in Fig. 14. The infected population explodes to a peak that is about 2.3 times the peak at which controls are on fully.

On the downward side of the controls, we release quarantine again when the infected population is about 1 per cent of the population. The results are plotted as a blue line, and shows that although there is no ignition to a large outbreak, the disease takes an increase such that it takes longer to reach a minimum.

## Appendix E: Comparison of SEAIR and SEIR predictions

Figure 16 shows a comparison of SEIR and SEAIR epidemic curve as a function of the fraction of asymptomatic infectious, *p*. Our simulations show that, when a low fraction of asymptomatic is considered (*p* = 15%), the SEIR overestimates the infectious count, and slightly underestimates the position of the peak. When a larger fraction is considered (*p* = 85%), the SEIR predictions fail with respect to both the size and timing of the peak.

**Figure 16:**
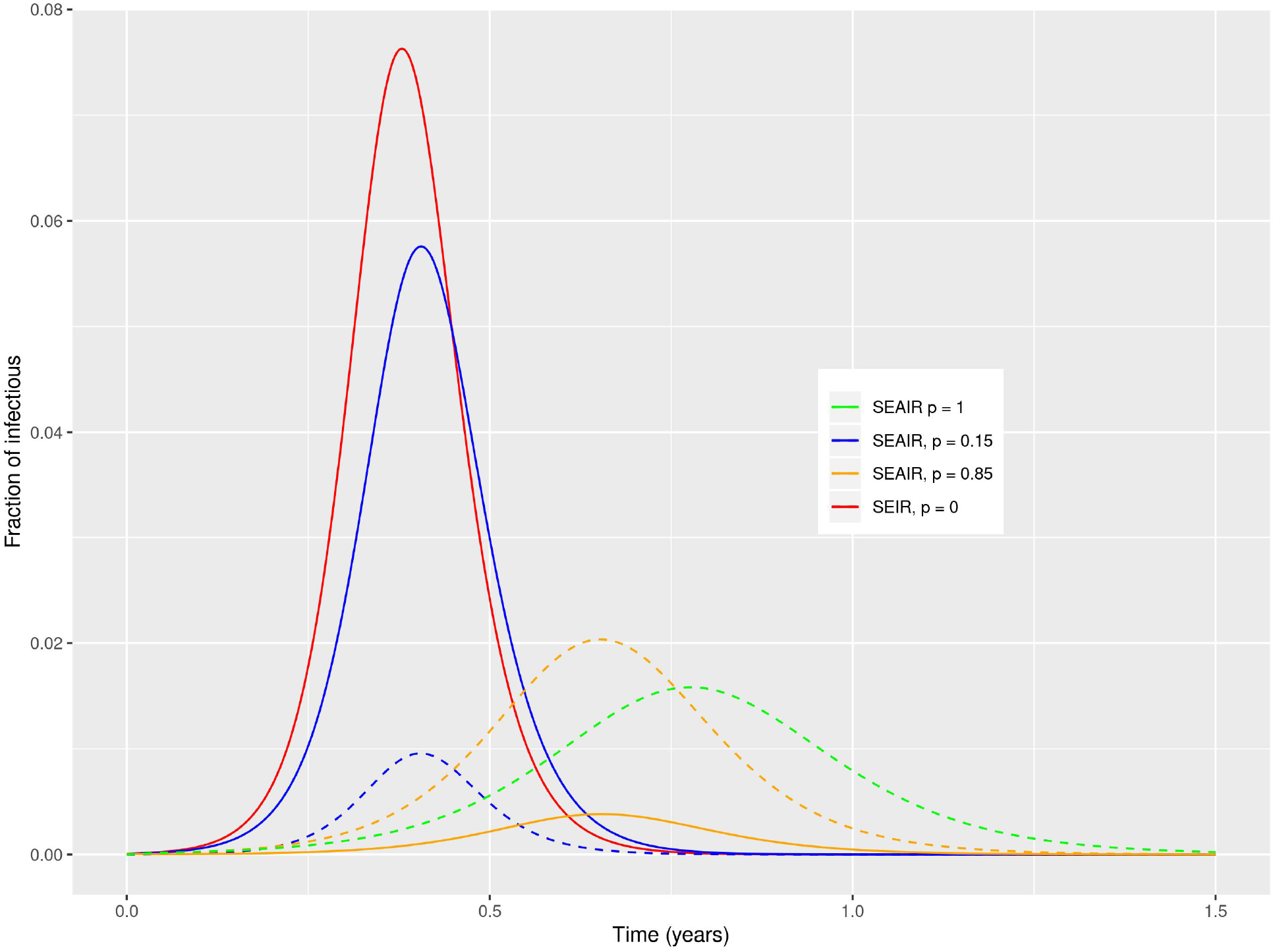
Comparison between SEIR and SEAIR for various fractions of the asymptomatic population, *p*. The solid lines represent the symptomatic infectious class, *I*. The dashed lines are the asymptomatic infectious, *A*. We observe that not including asymptomatic incorrectly estimates both size and timing of the infectious peak.

## Appendix F: Parameter values

The still evolving nature of the pandemic makes it generally challenging to estimate parameters accurately. The following table shows our choices for the parameters.

| Parameter | Value used<br>(years<sup>-1</sup>) | Range<br>(years<sup>-1</sup>) | Description | Reference |
| --- | --- | --- | --- | --- |
| $\beta_1$ | 53 – 107 | | Transmission rate, symptomatic infectious | Derived from $R_0$ |
| $\beta_2$ | 53 – 107 | | Transmission rate, asymptomatic infectious | Derived from $R_0$ |
| $\alpha$ | 36.5 | 32 - 182.5 | Disease incubation rate | [13] |
| p | 0.25 - 0.85 |  | Fraction of asymptomatic infectious | [4, 5] |
| $\gamma_A$ | 56 | 26 – 73 | Recovery rate, asymptomatic infectious | [3] |
| $\gamma_I$ | 52 | 26 – 73 | Recovery rate, symptomatic infectious | [3] |
| $\mu_d$ | 0.035 | 0.02 – 0.15 | Disease death rate | [14–17] |
| $\mu$ | 0.02 | | Background birth/death rate | |

## Appendix G: Numerical considerations regarding control

When social distancing is implemented as control, the contact rates are assumed to be such that *β*_*A*_ is a fraction of *β*_*I*_; i.e., we capture a fraction of the asymptomatic individuals. Typically, we assume that fraction is 50 per cent. When control is off, then the contact rates are equal, and set to values such that the value of *R*_0_ ≈ 2, which is our typical baseline. Control is released in a smooth fashion using an arctangent function to avoid discontinuities. This is reflected in the small deviations between control and release curves observed in the figures.

## Notes

### Competing Interest Statement

The authors have declared no competing interest.

### Funding Statement

BS acknowledges support from the NRL Base funding no. N0001420WX00410, as well
as the Office of Naval Research no. N00001419WX01322

